# Human brain single nucleus cell type enrichments in neurodegenerative diseases

**DOI:** 10.1101/2023.06.30.23292084

**Authors:** Chelsea X. Alvarado, Cory A. Weller, Nicholas Johnson, Hampton L. Leonard, Andrew B. Singleton, Xylena Reed, Cornelis Blauewendraat, Mike Nalls

## Abstract

Single cell RNA sequencing has opened a window into clarifying the complex underpinnings of disease, particularly in quantifying the relevance of tissue- and cell-type-specific gene expression. To identify the cell types and genes important to therapeutic target development across the neurodegenerative disease spectrum, we leveraged genome-wide association studies, recent single cell sequencing data, and bulk expression studies in a diverse series of brain region tissues. We were able to identify significant immune-related cell types in the brain across three major neurodegenerative diseases: Alzheimer’s Disease, Amyotrophic Lateral Sclerosis, and Parkinson’s Diseases. Subsequently, we identified the major role of 30 fine-mapped loci implicating seven genes in multiple neurodegenerative diseases and their pathogenesis.

## Introduction

Neurodegenerative diseases (NDD) encompass diseases characterized by progressive degeneration of cell types including neurons and glia in the central nervous system and/or peripheral nervous system. NDDs vary in their anatomic vulnerabilities and main affected neuropathologies, resulting in specific cell type dysregulation which may or may not be shared between NDDs.^1,2^ The complexity of biological processes and pathways involved in NDD pathogenesis has stymied progress in the understanding of disease and treatment development. While the expression of microglia in one cell type of the brain may be dysregulated, within the same disease state, other cell types may be regulated normally. Technology such as bulk RNA sequencing (RNA-seq) would be unlikely to identify these differences in expression since RNA-seq averages the expression of transcripts from all cells present in a tissue sample.

High throughput single-cell, or single-nucleus, mRNA sequencing (scRNA-seq, or snRNA-seq) technology provides a new window into understanding the functionally complex interactions between cell populations and across tissues. While bulk RNA-seq provides an assessment of expression averaged across a population of sampled cells, snRNA-seq allows for nuanced insight at the cell-type level. Instead of identifying gene set enrichment across an entire tissue, one can identify enrichment of gene expression partitioned by cell type, or changes in the composition of cell types themselves. uncertainty still limits our progress in better understanding the biological underpinnings of NDDs. In this report we leverage population-scale genome-wide association study (GWAS) data in conjunction with snRNA-seq in the brain to gain more specific insights into potential cell-type-specific mechanisms of risk within and across neurodegenerative diseases.

## Methods

### Single nucleus RNA-seq expression data

Data from adult human single nucleus brain tissues, consisting of the forebrain, midbrain, and hindbrain, was obtained from Siletti et al.^3^ The data was obtained by sequencing dissections from three healthy post-mortem donors. The subset of data used in this study consisted of snRNA-seq data. It included each of the 461 super clusters defined by Siletti et al. as being represented by the islands identified through a two-dimensional representation calculated by t-distributed stochastic neighbor embedding (t-SNE).^3^ Additionally, we calculated enrichment at the scRNA-seq data at the level of annotated cell type classes, using auto-annotation data provided by Siletti et al. in the supplementary materials of their manuscript.

### Genome-wide association study summary statistics

We included data from six NDD GWAS: Amyotrophic lateral sclerosis (ALS) from van Rheenen et al.^4^; Alzheimer’s disease (AD) from Bellenguez, et al.^5^; Frontotemporal lobar degeneration (FTLD) from Pottier, et al.^6^; Lewy body dementia (LBD) from Chia, et al.^7^; Parkinson’s disease (PD) from Nalls, et al.^8^; and Progressive supranuclear palsy (PSP) from Höglinger, et al.^9^ Data for ALS, AD, LBD, and PSP were obtained from GWAS Catalog (https://www.ebi.ac.uk/gwas) and data for FTLD and PD were obtained directly from the respective authors.

### MetaBrain eQTL summary statistics

We included expression quantitative trait loci (eQTL) summary statistics from MetaBrain, a large scale eQTL meta-analysis de Klein et al.^10^ For colocalization analysis, we included SNPs with a reported p-value no greater than 1×10^−4^ for the QTL component of the study.

### Cell Type Enrichment Analyses

We conducted cell type enrichment analyses using the R package MAGMA.^11,12^ Cell typing for each adult human brain snRNA-seq data and disease GWAS combination. Multiple testing correction (Bonferroni method) allows for efficient identification of enriched cell types. Required input data for conducting MAGMA analysis includes formatted GWAS summary statistics and a CellTypeDataset object (CTD).

Preprocessing of disease specific GWAS summary statistics and the creation of the CTD object, which holds cell specificity data, was performed using the R packages *MungeSumstats*^13^ and *ECWE*^14^ respectively. Quality control and munging of input data was conducted using methods available through the used R packages (see supplementary methods).

### Colocalization

We conducted Bayesian colocalization analysis using the R package *coloc*^15^ for all pairwise combinations of putatively significant autosomal NDD GWAS (p ≤ 5×10^−8^) and MetaBrain eQTLs (p ≤ 1×10^−4^). To account for the possibility of a single SNP influencing expression at multiple loci, we iterated over significant probes (see supplementary methods). We report associations with a posterior probability of at least 90%. A summary of data used and numbers of hits is included in **Supplementary Table S1**.

### Gene Expression Summarization

We summarized expression ranks for genes of interest within the single cell adult human brain transcriptome data set adult_human_20221007.loom from Siletti et al.^3^ Using custom R scripts we converted feature counts into TPM (transcripts per million). For a given sample, feature counts were divided by maximum nonredundant intron-removed exon lengths to correct for differences in gene length. Values were then multiplied by a sample-specific constant (10^6^ / T, where T is the sum of length-normalized counts) such that the resulting unitless vector sums to one million. We extracted exon lengths based on annotations from the GTF file used to originally annotate the single cell data (gb_pri_annot.gtf). We calculated the expression percentile rank for genes of interest using the empirical cumulative distribution function and then calculated the mean and median expression percentile rank (EPR) value for each gene for each tested cell type. In order to ease interpretation, we binned the EPR values into 3 classes: off (EPR < 10), low (10 < EPR < 90) or high (EPR > 90).

### Online methods

Additional methods details can be found in the Supplementary Materials section as well as in the Data and Code Availability section. All analyses comprising this workflow are summarized in **Figure 1**.

**Figure 1:**
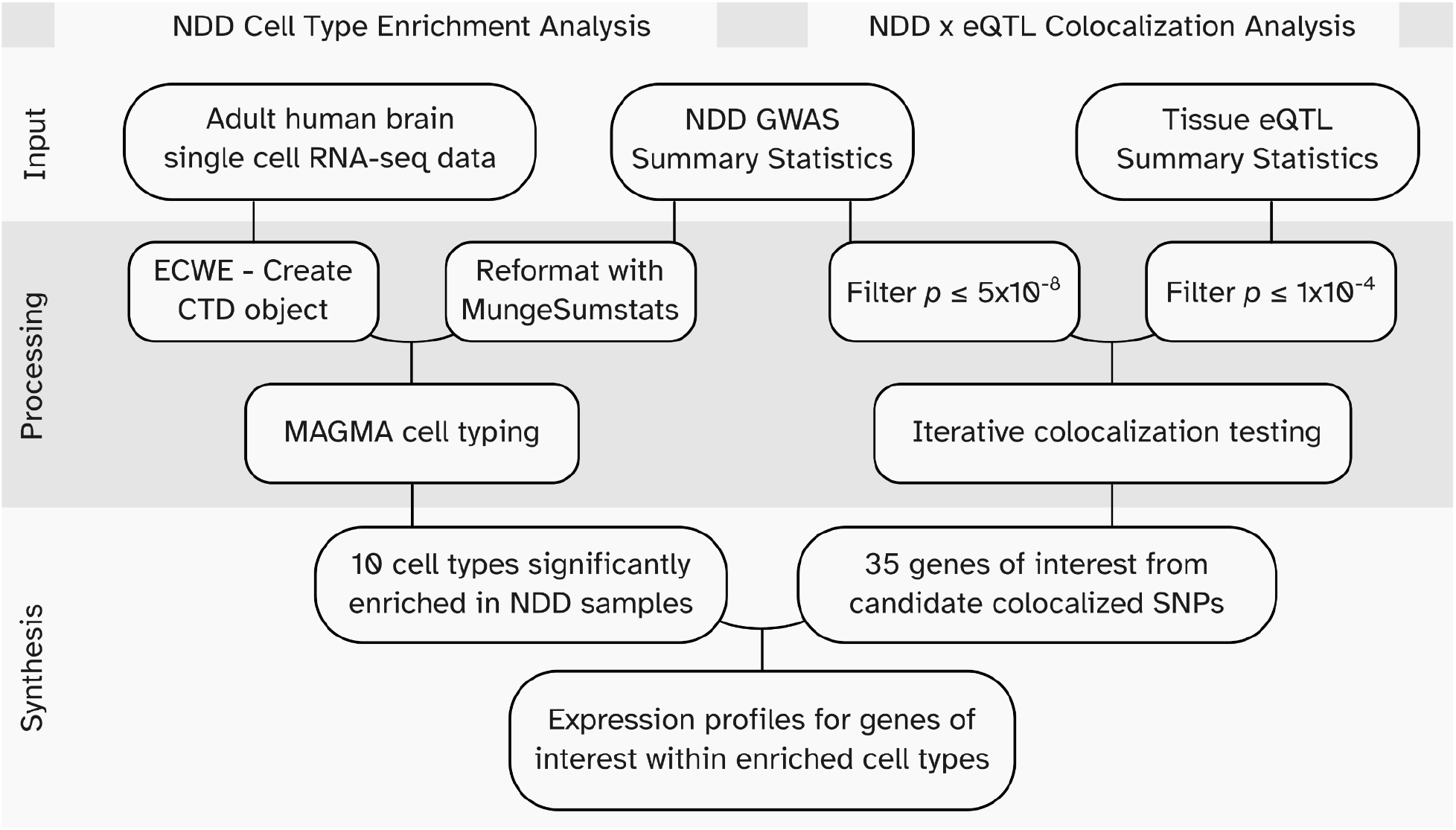
Workflow diagram. **NDD**: Neurodegenerative Disease. **GWAS**: Genome-Wide Association Study. **eQTL**: expression Quantitative Trait Loci. **ECWE**:Expression Weighted Cell Type Enrichment.^14^

## Results

### Cell type enrichments identified in AD, ALS, and PD

We identified significant cell type enrichments in the adult human brain for three out of six tested diseases: AD, ALS, and PD (**Table 1; Supplementary Table 2**). In general, we found that linear regression style enrichment analysis identified more significant enriched cell types than the top 10% enrichment style (n_linear_ = 30, n_Top 10%_ = 5). MAGMA. Celltyping documentation does state that using the linear regression enrichment mode results in more significant results due to overlapping cell type signatures.

**Table 1:** Significant Celltype enrichments across AD, ALS, and PD. Table provides information on each significantly enriched cell type after Bonferonni correction (p < 9.2×10^−4^). Results are provided for supercluster and class levels as well as differing enrichment modes. **CHRP ENDO**: Choroid Plexus and Endothelial. **ENDO**: Endothelial. **MAC**:Macrophage. **MGL**: Microglia. **MONO**: Monocyte. **NK**: Natural Killer cell. **OLIGO**: Oligodendrocyte. **OPC**: Oligodendrocyte precursor cell. **PER**: Pericyte. **TCELL**: T cell. **VSMC**: Vascular smooth muscle cell.

| Disease | Celltype | Genes | Beta | SE | P | Annotation Level | Enrichment Mode |
| --- | --- | --- | --- | --- | --- | --- | --- |
| AD | MAC | 16170 | 3.83E-03 | 0.00068234 | 9.82E-09 | Class | Linear |
| AD | Microglia | 16170 | 3.98E-03 | 0.000742 | 4.19E-08 | Supercluster | Linear |
| AD | MONO | 16170 | 3.36E-03 | 0.00064412 | 9.25E-08 | Class | Linear |
| AD | Microglia | 1585 | 1.28E-01 | 0.027081 | 1.19E-06 | Supercluster | Top 10% |
| PD | MONO | 16178 | 0.0031777 | 0.00067611 | 1.31E-06 | Class | Linear |
| AD | MGL | 16170 | 3.56E-03 | 0.00075854 | 1.35E-06 | Class | Linear |
| PD | TCELL | 16178 | 0.0032981 | 0.00071908 | 2.27E-06 | Class | Linear |
| PD | CHRP ENDO | 16178 | 0.0028723 | 0.00065541 | 5.92E-06 | Class | Linear |
| PD | MAC | 16178 | 0.0031491 | 0.00071891 | 5.97E-06 | Class | Linear |
| PD | NK | 16178 | 0.002998 | 0.00069001 | 7.02E-06 | Class | Linear |
| PD | Thalamic excitatory | 16178 | 0.0032777 | 0.00078414 | 1.47E-05 | Supercluster | Linear |
| PD | PER | 16178 | 0.0030213 | 0.00072459 | 1.54E-05 | Class | Linear |
| PD | CGE interneuron | 16178 | 0.0032385 | 0.00078432 | 1.83E-05 | Supercluster | Linear |
| PD | LAMP5 LHX6 and Chandelier | 16178 | 0.0032582 | 0.00079491 | 2.09E-05 | Supercluster | Linear |
| PD | Midbrain derived inhibitory | 16178 | 0.0032348 | 0.00079837 | 2.56E-05 | Supercluster | Linear |
| PD | Eccentric medium spiny neuron | 16178 | 0.0031299 | 0.00077946 | 2.98E-05 | Supercluster | Linear |
| PD | VSMC | 16178 | 0.0028347 | 0.00071616 | 3.80E-05 | Class | Linear |
| PD | OLIGO | 16178 | 0.0033272 | 0.00084917 | 4.48E-05 | Class | Linear |
| AD | NK | 16170 | 2.51E-03 | 0.00065687 | 6.65E-05 | Class | Linear |
| PD | Hippocampal dentate gyrus | 16178 | 0.0029692 | 0.00078409 | 7.66E-05 | Supercluster | Linear |
| PD | OPC | 16178 | 0.0031794 | 0.00085042 | 9.29E-05 | Class | Linear |
| PD | Upper rhombic lip | 16178 | 0.0030704 | 0.00082409 | 9.78E-05 | Supercluster | Linear |
| AD | MGL | 1080 | 1.16E-01 | 0.031771 | 1.27E-04 | Class | Top 10% |
| AD | NK | 856 | 1.30E-01 | 0.035634 | 1.35E-04 | Class | Top 10% |
| AD | MAC | 1073 | 1.16E-01 | 0.031778 | 1.37E-04 | Class | Top 10% |
| PD | ENDO | 16178 | 0.002692 | 0.00074809 | 1.61E-04 | Class | Linear |
| PD | Hippocampal CA1 3 | 16178 | 0.0027432 | 0.00080036 | 3.06E-04 | Supercluster | Linear |
| PD | MGE interneuron | 16178 | 0.0026543 | 0.00078158 | 3.43E-04 | Supercluster | Linear |
| PD | Deep layer intratelencephalic | 16178 | 0.0026773 | 0.00079243 | 3.65E-04 | Supercluster | Linear |
| PD | Oligodendrocyte precursor | 16178 | 0.0028421 | 0.00084173 | 3.68E-04 | Supercluster | Linear |
| PD | Mammillary body | 16178 | 0.0027659 | 0.00082975 | 4.30E-04 | Supercluster | Linear |
| PD | Upper layer intratelencephalic | 16178 | 0.0026419 | 0.00079874 | 4.72E-04 | Supercluster | Linear |
| PD | Committed oligodendrocyte precursor | 16178 | 0.0024936 | 0.00077006 | 6.03E-04 | Supercluster | Linear |
| AD | MONO | 972 | 1.05E-01 | 0.03294 | 7.06E-04 | Class | Top 10% |
| PD | Bergmann glia | 16178 | 0.0024558 | 0.00078146 | 8.39E-04 | Supercluster | Linear |
| ALS | MONO | 16094 | 1.78E-03 | 0.00056656 | 0.00085158 | Class | Linear |

In PD, we ran MAGMA cell typing analysis on three variations/subsets of Nalls et al. meta-GWAS^8^: the complete meta-GWAS; meta-GWAS excluding 23andMe data; and meta-GWAS excluding 23andMe and UK Biobank data. In the first two variations tested, no cell types at the supercluster or class level reached significant enrichment after Bonferroni correction (p < 9.2×10^−4^). The only GWAS variation that identified significant cell type enrichments was when the full PD meta-GWAS was used in the cell typing analysis. We identified 14 enriched cell types when using linear regression analysis at the supercluster level. At the class level, we identified 10 enriched cell types (**Supplementary Table S3**). The top enriched cell type at the supercluster and class level are *thalamic excitatory* and *monocytes* (p_thalmic excitatory_ = 1.467 ×10^−5^, *β*_thalmic_excitatory_ = 3.278×10^−3^, p_monocytes_ = 1.314×10^−6^, *β*_monocytes_ = 3.178×10^−3^), with monocytes being the most significantly enriched cell type in PD. Genes enriched in the monocyte cell type include *FCN1, CLEC12A, S100A4, TNFRSF1B, IFI30, LYZ, CYTIP, FGR, LILRB2, KYNU (***Supplementary Tables S4-5**).

In AD, we found microglia to be the only significantly enriched cell type at the supercluster level in both linear and top 10% analyses (p < 9.2×10^−4^ ; p_linear_ = 4.193×10^−8^, *β*_linear_ = 3.978×10^−3^, *β* p_Top 10%_ = 1.195×10^−6^, *β*_Top 10%_ = 0.1278) for risk loci. At the class level, four cell types were found to be significant after Bonferroni correction and in both analyses. Significantly enriched cell types identified include macrophages, monocytes, microglia, and natural killer cells (**Supplementary Table S6**). AD was the only disease to have significant enrichments when using the *Top 10%* enrichment analyses.

In ALS, we identified one significant cell type enrichment using linear regression enrichment analysis and at the class level annotations. *Monocytes* were the only cell type to reach significance (Bonferroni-corrected threshold of p < 9.2×10^−4^); p_Monocytes_ = 8.516×10^−4^, *β*_monocytes_ = 1.778×10^−3^; **Supplementary Table S7**). We did not detect significant cell type enrichments in FTLD, LBD, and PSP. No cell types at either the supercluster or class level reached significance after MAGMA-implemented Bonferroni correction (p < 9.2×10^−4^; **Supplementary Table S2, Supplementary Tables S8-10**).

### Colocalization

Across all diseases tested we fine mapped a total of 205 association signals at posterior probability > 90%. This included 89 unique genes identified as harboring putative causal associations. Of these 205 associations, 20 were centered around the *HLA* region, with colocalized omic associations in the cerebellum and cortex (the latter in multiple ancestry groups) suggesting extremely complicated risk of neuroinflammation in this part of the genome. Interestingly, *GRN* is fine-mapped using colocalized expression QTL (eQTL) signals in the cerebellum for AD and the cortex for PD suggesting related but potentially different mechanisms. The *TMEM175*/*GAK* region shows multiple colocalized signals across multiple diseases (PD and LBD). Of particular interest is that *GAK* and *TMEM175* both are fine-mapped to the same SNP (rs6599388) with eQTL effects in opposite directions in LBD. At the same time, regional signals for the gene *SLC26A1* were also fine-mapped for PD, LBD and AD with the effect in PD being detected in the spinal cord while the other disease QTLs were localized to the cortex.

Variants exhibiting a colocalization posterior probability ≥ 90% are summarized in **Figure 2** and **Table 2**. Extended data is available in **Supplementary Table S11**. 30 loci were fine-mapped to a single gene per disease by leveraging QTL data (see **Table 2** for details^10,16^). Of these, six are known druggable genes including *ABCA1, ADAM10, CD55, FGF7, OXGR1* and *POLE* (see **Supplementary Table S12**). Mining additional data on these druggable genes from the omicSynth^16^ database (**Supplementary Table S13**), we note that *ABCA1* has putative functional multi-omic associations with AD and PSP in blood and brain tissues. *ADAM10* has a similar pattern of functional inferences in AD. *CD55* is shown to have multiple significant functional inferences in brain tissues mediating risk for AD and LBD. However, *FGF7* does not display any significant functional inferences in any diseases from the database query. Methylation QTLs in blood connect PSP and FTD at *OXGR1* via functional inferences using SMR, while multiple brain, blood and nerve associations connect this gene with PD risk across both expression and methylation QTLs. A similar pattern of disease and tissue associations is seen for *POLE*, although there is no significant neural tissue association for PD and PSP is also connected via blood eQTLs to the same gene. Of the fine mapped loci, 22 (Supplemental Table S16) have also been nominated elsewhere as potential therapeutic targets with likely functional impacts on neurodegenerative disease risk in the context of methylation, expression, protein or chromatin QTLs detailed in the omicSynth web application.^16^

**Figure 2:**
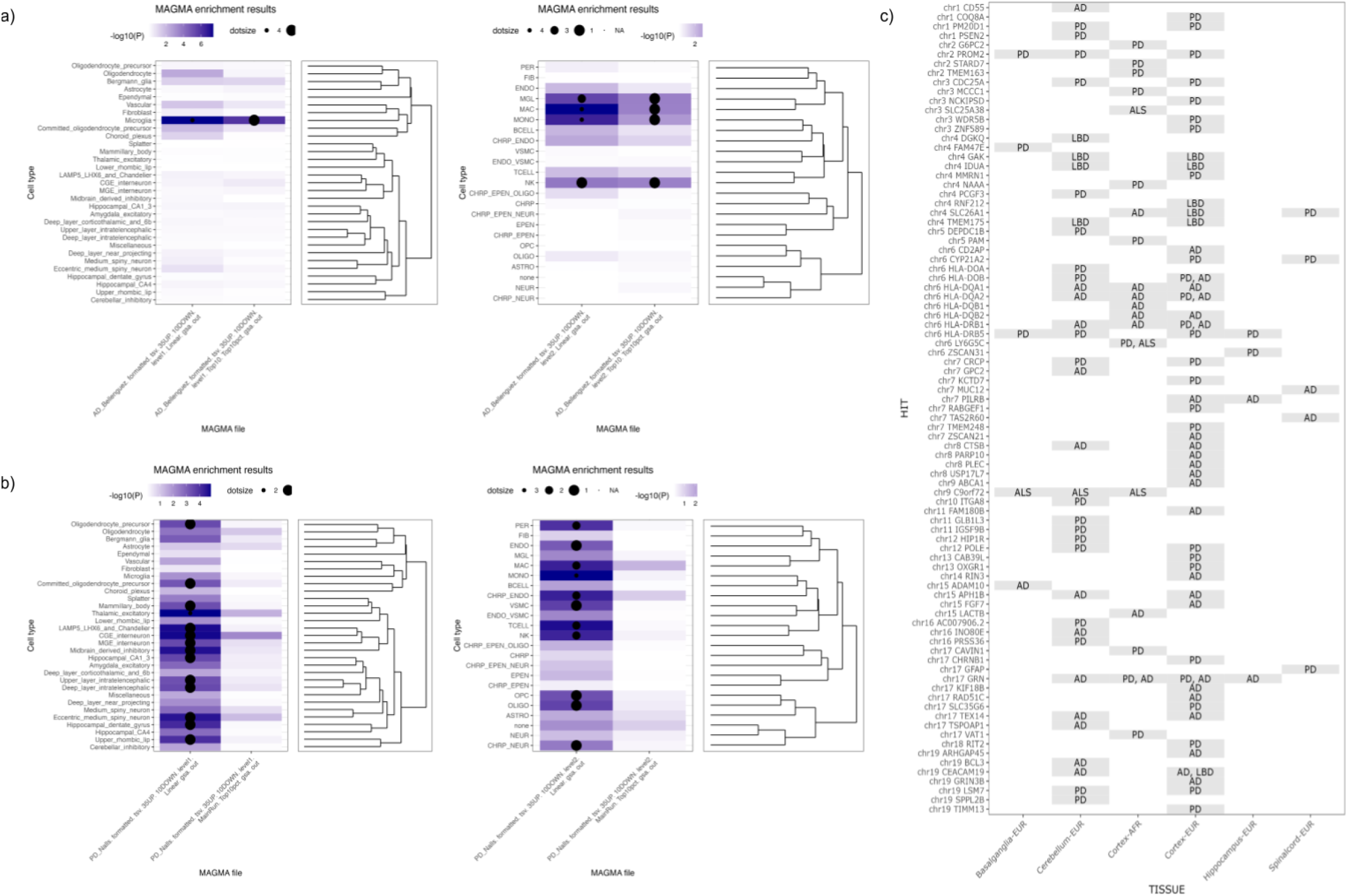
Graphical summaries of cell type enrichment and colocalization results. **A**. Tile plot comparison of significant cell type enrichments, by disease at each level for both MAGMA.Celltyping analysis. A Bonferonni significance line is provided on the bar chart portion of the image. **B**. Comparison of significant cell type enrichments, with celltype dendrogram, by disease at the class level for both MAGMA.Celltyping analysis. **C**. Summary of fine-mapped loci per disease. AD = Alzheimer’s Disease, ALS= Amyotrophic Lateral Sclerosis, PD = Parkinson’s Disease.

**Table 2:**
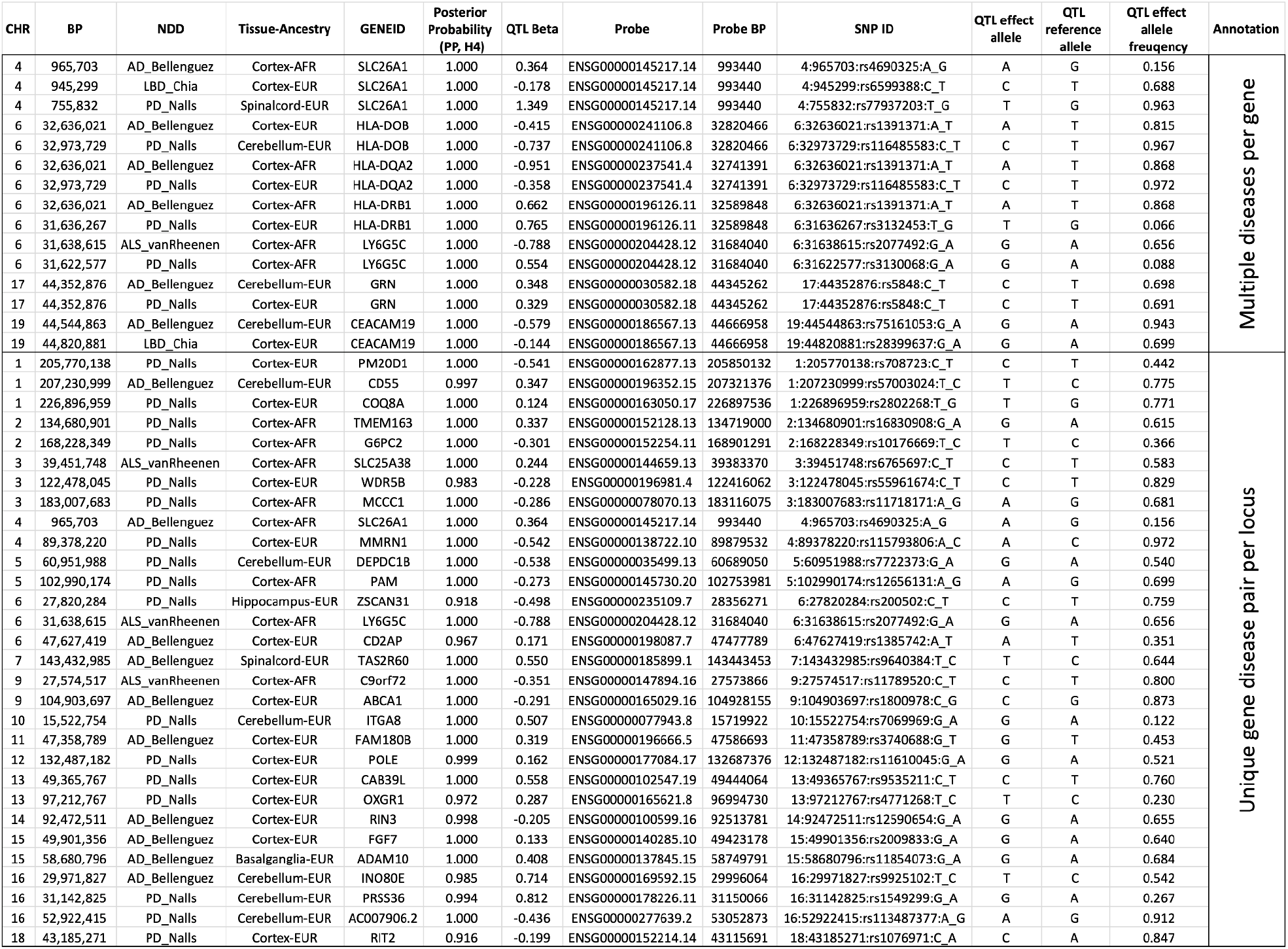
Summary of colocalized signals. This summary details all associations suggesting a single gene associated with multiple diseases of interest or a single gene per locus at posterior probability above 90%.

### Cell type resolution of colocalized genes

We further evaluated gene expression from snRNA-seq used in enrichment analysis. We calculated the mean and median expression percentile rank (EPR) for each gene across cells corresponding to the nominated supercluster cell types identified in our enrichment analyses (**Table 1**) and compared the aggregate mean and median values against the nominated colocalized genes (**Supplementary Figure S2, Supplementary Tables S15-16**). In order to ease interpretation we binned the mean EPR values into three categories based on the mean EPR value for each gene-cell type combination: *off, low*, and *high* (see methods).

We identified 10 colocalized genes with *high* median EPR values out of the 14 tested genes. The gene *PAM* has 6 cell type combinations (CGE interneuron, Eccentric medium spiny neuron, MGE interneuron, Hippocampal dentate gyrus, Thalamic excitatory, and Mammillary body) classified as *high* (**Figure 3**). Overall, the Mamillary body cell type had the greatest count of eight *high* EPR genes.

**Figure 3:**
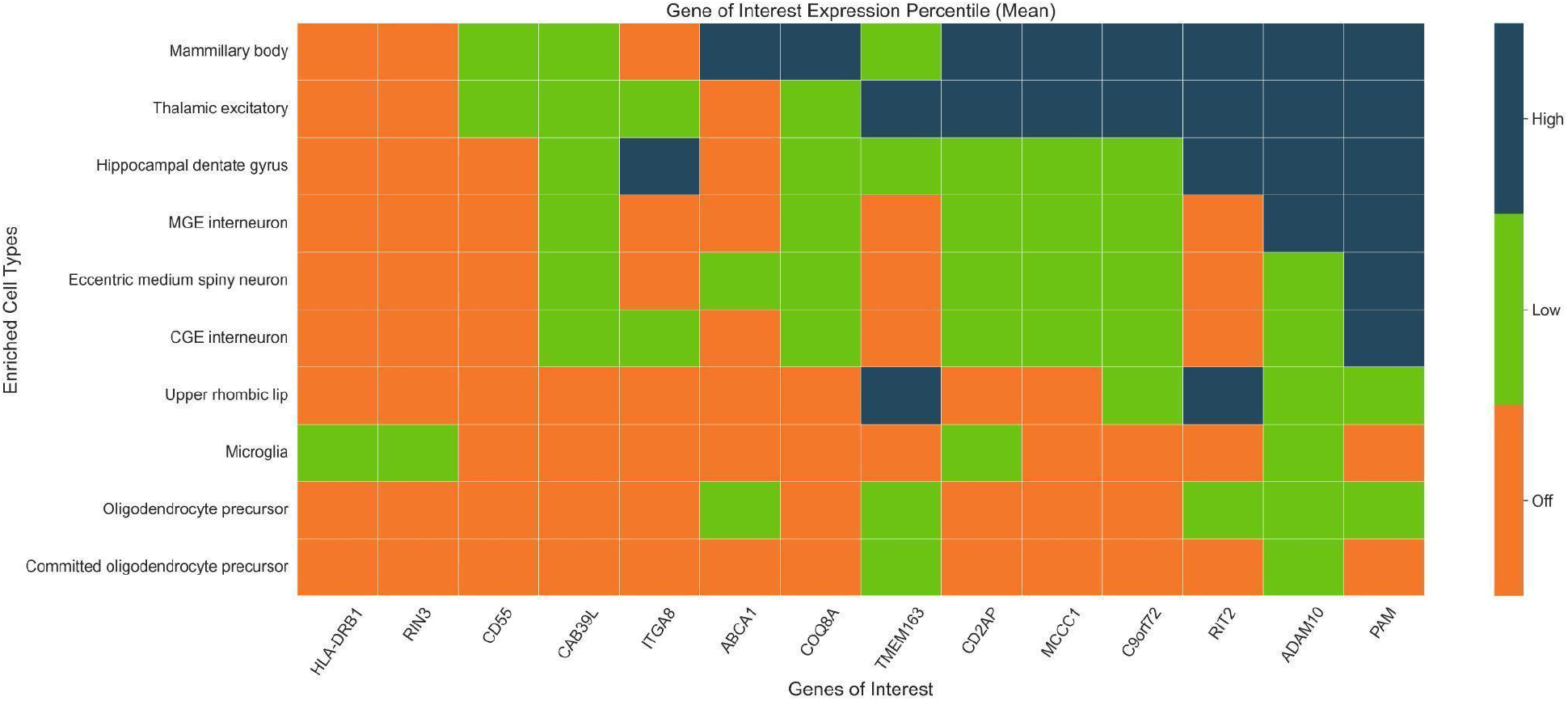
scRNA-seq expression distribution across colocalized genes. This graphical representation visualizes the scRNA-seq expression distribution across MAGMA nominated enriched supercluster cell types. Expression percentile rank (EPR) for each cell-gene combination was binned into *high expression* (dark blue, median expression within top 10% of all genes), *low expression* (green, middle 80%) or *off* (orange, bottom 10%).

## Discussion

We identified enriched cell types in various brain regions for three (AD, ALS, and PD) out of six tested NDDs utilizing snRNA-seq data of the 461 superclusters identified by Siletti et al.^3^ We did not detect significant cell type enrichments, at either the supercluster or class level, in FTLD, LBD, and PSP. We speculate that due to the smaller sample sizes of each GWAS we were unable to identify any significantly enriched cell types after applying the MAGMA-implemented Bonferroni correction (p < 9.2×10^−4^; **Supplementary Table S2, Supplementary Tables S8-10**).

All significantly enriched cell types had an associated positive beta for risk genes, and our identified significant cell type enrichments fall in line with current knowledge on cell types implicated in various NDD pathologies which we will discuss further. Broadly, our results highlight the importance of immune-related cell types in the pathology of varying NDDs. Additionally, our results add to the growing evidence that NDDs, such as AD, ALS, and PD, are highly related to autoimmune diseases or may even be autoimmune diseases of the brain.^17–20^

Monocytes were the only cell type significantly enriched across the three diseases AD, ALS, and PD. It is the most significantly enriched cell type in ALS and PD (p_ALS_=8.51×10^−4^, p_PD_ = 1.31×10^−6^) and the third most enriched cell in AD (p_AD_ = 9.25×10^−8^). Monocytes are the precursor cells to dendritic cells (DCs) and macrophages which both play important roles in immune response, neuroinflammation, and neuroimmunological response.^21–23^ Of the genes enriched in the monocyte cell type (previously described in our results), *KYNU* is a gene of note, being part of the tryptophan metabolic pathway which has been found to play a role in Aβ formation.^19^

Microglia are another significantly enriched cell type identified in our analyses, though only significantly enriched in AD. Previous literature highlights the role that microglia play in neuronal loss and neuroinflammation as well as their involvement in immune response.^24–26^ Microglia are known to function similarly to DCs and macrophages, derived from monocytes.^25^ In AD literature, microglia is implicated as an affected cell type associated with neuroinflammation.^25,27^ Our results highlight microglia as the second most enriched cell type in AD after Bonferroni correction (p_AD_ = 4.19×10^−8^). Microglia were found to be nominally significant in LBD and PD (p_LBD(Linear)_ = 0.0119, p_PD(Linear)_ = 9.25×10^−3^, p_PD(Linear)_ =1.96×10^−03^, **Supplementary Tables 2, 3, 6**). Interestingly, while not shown as enriched in our ALS analyses, microglia have been shown to play an important role in the pathogenesis of ALS in part due to their activation being neurotoxic to motor neurons.^25,27–30^ It is possible that due to the limited sample size in the used snRNA-seq data, the analyses were unable to detect any significant cell type enrichment in microglia for ALS.

Common risk factors across neurodegenerative diseases have always been of interest to the basic science and therapeutic industries. The association at the *SLC26A1* locus is of particular interest across multiple NDDs, as it is strongly associated with *IDUA* protein level in QTL studies, with deficiencies in this protein causing severe lysosomal storage disorders.^31,32^ The *HLA* region is a complex locus from a structural genetic standpoint, but also in terms of general risk of neuroinflammation so not surprisingly resulting in a number of fine-mapped associations across this locus for multiple genes.^5,33^ *GRN* is a positive control for this colocalization analysis effort as previous efforts show increased genetic risk in the region coincides with decreased expression of the gene in PD, AD and ALS.^34^

Fine-mapping efforts localizing signals to single genes within risk loci help to identify novel therapeutic targets with known biological plausibility. *ABCA1* is known to be associated with Tangier disease, which is a rare autosomal recessive disorder with low plasma levels of high-density lipoprotein (HDL) causing peripheral neuropathy.^35^ *ADAM10* is implicated in the formation of amyloid plaques in the brain and the processing of APP.^35,36^ CD55 is known to interact with viruses and cause neuroinflammation potentially leading to increased AD risk.^37^ *COQ8A* mutations have been shown to cause coenzyme Q10 deficiency leading to autosomal recessive ataxia, cerebellar atrophy, and progressive movement disorders.^38^ *C9orf72*, known to be linked to ALS, acts as a positive control here and displays a unique ALS/FTD colocalization. *ADAM10* and *C9orf72* were also nominated as potential therapeutic drug targets for neurodegenerative disease via Mendelian randomization.^16^

Limitations to this study generally relate to availability of data in this context. First and foremost, there is a limited amount of multi-ancestry or non-European data available for the GWAS and single-cell or QTL resources used here. This potentially introduces bias into therapeutic development and precision medicine applications. Secondly, low sample sizes for single nucleus analyses (in terms of the number of humans involved) reduces our ability to generate eQTL databases as compared to coarse methods of bulk RNA sequencing at scale.

Here we provided insights that could potentially aid in therapeutic development for NDDs. On the macro-level, we have identified cell-type level enrichments associated with disease risk in multiple neurodegenerative diseases allowing biologists and drug developers to better focus their mechanistic and therapeutic research. On the micro-level, we have used eQTL colocalization methods to narrow down the large tracts of associated loci in GWAS to potentially functional variants, metaphorically going from a neighborhood to building level resolution on a map.

## Data Availability

All GWAS summary statistics used in this report are available at https://nih-card-ndd-smr-home-syboky.streamlit.app/About
Single nucleus (sn) RNA-seq expression data is available at https://github.com/linnarsson-lab/adult-human-brain which is the repository associated with https://doi.org/10.1101/2022.10.12.511898
eQTL data is available at the MetaBrain browser: https://www.metabrain.nl/
Code used for analysis is available at https://github.com/NIH-CARD/NDD_Single_Cell

https://nih-card-ndd-smr-home-syboky.streamlit.app/About

https://github.com/linnarsson-lab/adult-human-brain

https://www.metabrain.nl/

## Data and Code availability

Code used to generate and process our data can be accessed at our Github Repository. All data for this project are publicly available via the original publications accessed by our team. Summaries of enrichment and colocalization data are also available to browse in our community target discovery and due diligence resource omicSynth web application.^16^

