## Supplemental Materials for "Human brain single nucleus cell type enrichments in neurodegenerative diseases"

### Supplementary Materials

##### *Preprocessing: MAGMA.Celltyping*

Preprocessing on the single cell data for use with MAGMA.Celltyping was primarily conducted using the MungeSumstats and EWCE R packages available through bioconductor [1-3[1–3]]. The workflow to preprocess GWAS and single cell expression data is detailed in multi-marker analysis of genomic annotation (MAGMA) Cell typing documentation. The first step was to ensure GWAS summary statistics were in the appropriate format for use with MAGMA Cell typing by using the MungeSumstats package method *format_sumstats*. All GWAS summary statistics used were already on or lifted to build GRCh38 during preprocessing through MungeSumstats built-in methods. MungeSumstats standardizes GWAS summary statistics and conducts quality control steps when provided with the minimum SNP ID, chromosome, and base pair position in the first three columns and at least one signed/effect column (Z, OR, BETA, LOG_ODDS, SIGNED_SUMSTAT). In order to compare GWAS summary statistics to a gene-level transcriptomic cell-type reference, the GWAS data must be converted to a gene-level signature using a method provided by the MAGMA.Celltyping package, *map_snps_to_genes*.

The second step is creating the CellTypeDataset object (CTD) using scRNA-seq data in order to obtain gene signatures for cell types. EWCE calculates cell type specificity at two user specified annotation levels in order to create the CTD object. Before calculation of specificities we removed uninformative genes (expressed sporadically) using the function, *drop.uninformative.genes.* The main method used to create the object was *generate_celltype_data*. The original expression data is available in multiple formats through the Linnarson Lab (github). We obtained the supercluster level data as a loomfile. The first instance of each gene in the expression matrix was kept and duplicates were removed. Metadata for each supercluster and class was obtained from the supplementary materials on the original preprint manuscript (preprint).

The single cell RNA-seq data expression profiles of brain cell types identified in Siletti., et al. [4[4]], can be compared against disease loci to identify affected cell types. In order to calculate any significant cell type enrichment associations for each disease we used the MAGMA Celltyping R package, which is available through bioconductor. We used our preprocessed GWAS summary statistics for each disease and the CTD object generated in preprocessing in order to calculate enrichment. We chose to run both cell type association analyses available (Linear and Top 10%) which correspond to the association tests available in general MAGMA analysis [5[5]]. The package, by default, conducts a Bonferroni correction on the results which we utilized to look across diseases in order to draw conclusions. Additionally, results of the association analysis are returned for the first and second level annotations corresponding to the supercluster type and class type.

##### *Metabrain eQTLs*

The data includes putative cis-eQTLs, *i.e.* SNPs associated with modular expression of local genes as measured by hybridization with probes targeting genomic regions within one megabase of the SNP. The data primarily cover a population of European ancestry, and includes measurements for the basal ganglia, cerebellum, cortex, hippocampus, and spinal cord. African ancestry eQTLs were available for the cortex. We retrieved putatively significant cis-eQTLs from the MetaBrain besd files using *smr* v1.3.1, Zhu et al. (2016) with the options --descriptive-cis --beqtl-summary --query 1e-4.[6,7]

##### *Colocalization*

We conducted Bayesian colocalization analysis using coloc.abf (from *coloc* v5.1.0.1) in *R* (v4.2.2). Because *coloc* is predicated on analysis being conducted on a genomic region, as opposed to whole chromosomes,[8] we usd a computational approach to detect local clusters of putative significant disease GWAS hits. Briefly, we iterated over potential cM threshold values ($D$) within the interval (0.01, 1) with steps of 0.01 cM. Map unit estimates were derived from linear interpolation of recombination rates reported by Loh et al.[9] For every value $D$, SNPs with $p \leq 5\text{x}10^{-8}$ were added to the adjacent cluster if the distance $\leq D$ cM. We evaluated the decay of genome-wide clusters $N_D$. as a function of threshold $D$, selecting the lowest threshold per GWAS that achieved at least 90% effective reduction in cluster count (*i.e.* the minimum $D$ where $[N_{0.01} - N_D] / [N_{0.01}-N_1] > 90\%$). After selecting this threshold $D$ (Supplementary Figure S1), we extended cluster boundaries by an additional $D/2$ in both directions and performed separate colocalization analysis on eQTL and NDD GWAS data within the intervals. The eQTL allele frequency and $\beta$ estimates were harmonized to match the NDD GWAS A1 and A2 values. We filtered by an eQTL minimum p-value of $1\text{x}10^{-4}$, and a minimum NDD GWAS p-value of $5\text{x}10^{-8}$. Data were not otherwise LD-pruned or filtered due to MAF. For each defined window we called the *coloc.abf* function, supplying $\beta$ values directly, var($\beta$) as the square of standard error, and sdY=1 for the eQTLs as expression data had been normalized. Other parameters retained default values.

Individual base pair positions per chromosome must be unique to run *coloc*. One approach to satisfying the uniqueness requirement is selecting the most significant expression probe per SNP. However, the top probe approach is problematic if we wish to detect SNPs that influence expression at multiple loci. To circumvent this problem, we used an iterative colocalization approach. We began by testing for colocalization with the most significant expression probe (defined by lowest p-value) per SNP, and recorded any associations with a resulting posterior probability ≥ 90%. We then modified the eQTL data to include the second most significant expression probe, after removing the ‘hits’ from the previous step. We repeated this test-record-remove pattern until no association with posterior probability ≥ 90% was found.

### Supplementary Figures

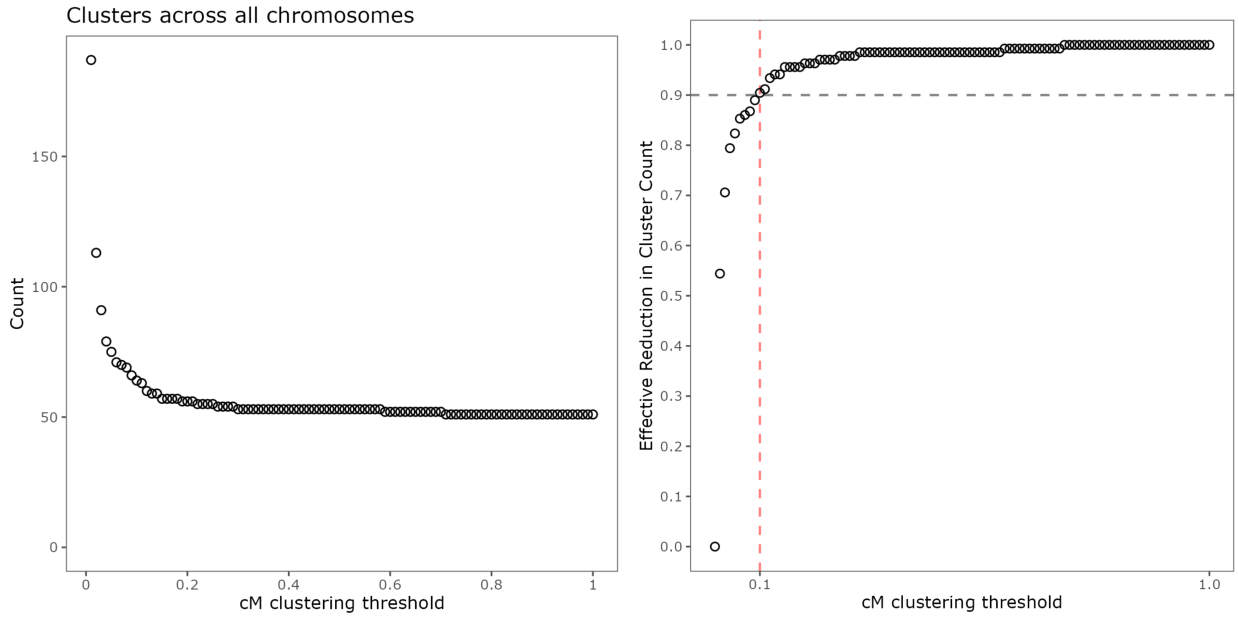


**Supplementary Figure S1**. SNP Clustering. Determination of SNP clustering cM thresholds. We chose $\pm$0.1 cM (red dashed line) as our clustering threshold, which achieved a 90% reduction in cluster counts.

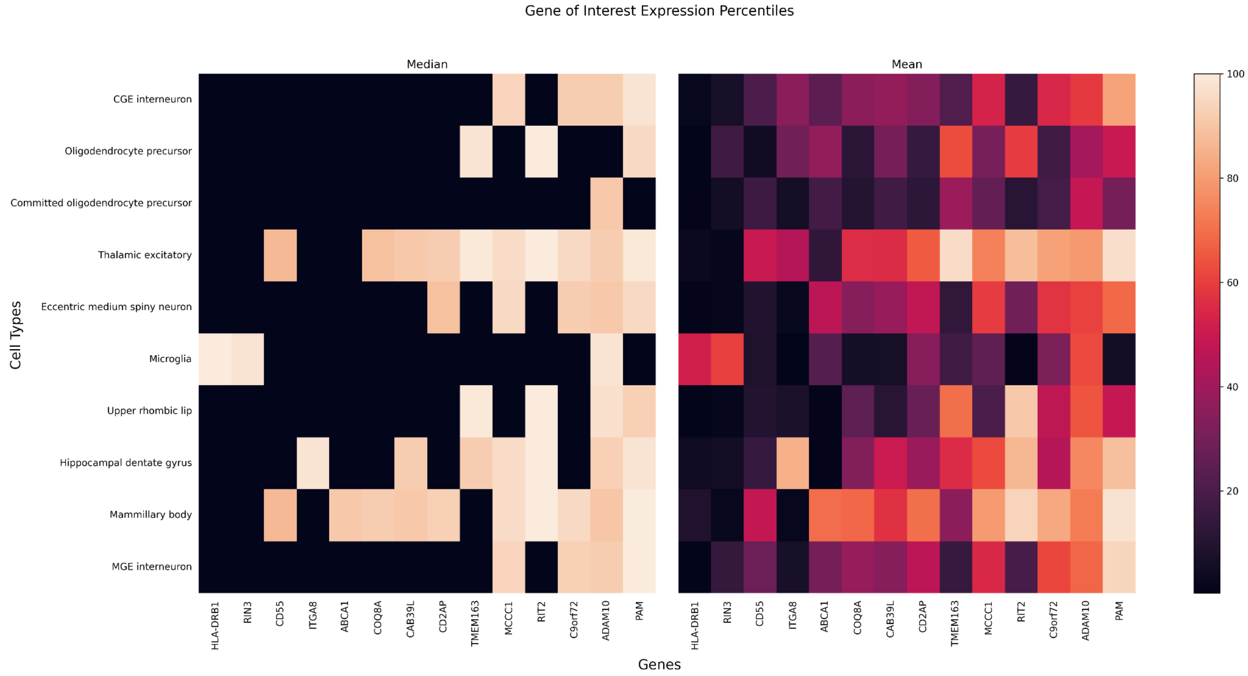


**Supplementary Figure S2**: scRNA-seq expression for colocalized gene hits. We calculated the expression percentile rank (EPR) for genes of interest using the empirical cumulative distribution function and then calculated the mean and median EPR value for each gene across significantly enriched disease relevant cell types.

#### Supplementary Tables

Tables are included as individual tabs within a separate excel (.xlsx) document. Captions are provided below:

**Supplementary Table S1**: Colocalization Summary Statistics. **Top**: Input data used for colocalization analysis. **Middle**: Counts for unique SNP colocalization associations between NDD and eQTL data sets.. **Bottom**: Counts for unique gene colocalization associations between NDD and eQTL data sets.
**Supplementary Table S2**: MAGMA.Celltyping Results. This table provides all MAGMA analyses results regardless of significance and for all combinations of the six NDD and two different enrichment analyses modes. The number of genes used for each analysis is provided. The number of tested genes varies only when using Top % enrichment analysis since it only uses the 10% most cell-type-specific genes to calculate enrichment.
**Supplementary Table S3**: PD MAGMA.Celltyping results. This table provides all MAGMA analyses results for Parkinson's disease and regardless of significance.
**Supplementary Table S4**: Enriched genes by cell type. This table provides the number of genes enriched in each cell type, as well as the gene symbols for each enriched gene. Enriched genes are provided for each unique cell type at both the supercluster and class levels. The annotation level of each cell type is provided.
**Supplementary Table S5:** Additional supercluster metadata. This table provides select metadata for each of the 461 clusters identified by Siletti., et al. Included are top enriched genes, top three regions, and top three dissections for each supercluster. Additional metadata can be found in the supplementary materials associated with Siletti., et al's manuscript.
**Supplementary Table S6**: AD MAGMA.Celltyping results. This table provides all MAGMA analyses results for Alzheimer's disease and regardless of significance.
**Supplementary Table S7**: ALS MAGMA.Celltyping results. This table provides all MAGMA analyses results for Amyotrophic Lateral Sclerosis and regardless of significance.
**Supplementary Table S8**: LBD MAGMA.Celltyping results. This table provides all MAGMA analyses results for Lewy Body Dementia and regardless of significance.
**Supplementary Table S9**: FTLD MAGMA.Celltyping results. This table provides all MAGMA analyses results for Frontotemporal Dementia Lobar Degeneration and regardless of significance.
**Supplementary Table S10**: PSP MAGMA.Celltyping results. This table provides all MAGMA analyses results for Progressive Supranuclear Palsy and regardless of significance.
**Supplementary Table S11**: Extended Colocalization Results. Includes all associations between NDD GWAS and eQTL colocalization testing with posterior probability of hypothesis 4 (PPH4) of at least 90%.
**Supplementary Table S12**: Colocalization Drugs. Colocalization hits with posterior probability of hypothesis 4 (PPH4) were compared against existing data on therapeutics from Finan et al (2017) and the Drug Gene Interaction Database. Data provided includes the gene symbol, source of the interaction claim, drug claim name, primary drug claim name, ENSG ID, druggability tier,

chromosome, position information, strand, number of GWAS linkage disequilibrium intervals the gene overlaps, small molecule druggability status of a gene's protein products, biotherapeutic druggability status of a gene's protein products, if a gene's protein products are involved in the absorption, distribution, metabolism, and excretion (ADME) of a compound, and the long name of the gene. Data, if available, is provided for gene claim names, interaction type with therapeutic molecule, drug names, CHEMBLID, interaction group and PMIDS.
**Supplementary Table S13**: Colocalization SMR Hits. Cross reference of colocalization identified genes to SMR associations from Alvarado et al. (2023). Data is provided on the SMR tissue tested and in which neurodegenerative disease it was tested. Standard summary statistical data is provided including probe and SNP chromosome and base pair position, effect and alternative alleles, beta, standard error, p value of SMR analysis, and the HEIDI score. More information on SMR result interpretation can be found here.
**Supplementary Table S14**: Intersection of our colocalization analysis and loci implicated via omicSynth.[10]
**Supplementary Table S15**: Mean Expression Percentile Ranks for scRNA-seq expression distribution across colocalized genes.
**Supplementary Table S16**: Median Expression Percentile Ranks for scRNA-seq expression distribution across colocalized genes.
